# Association of Immune-Related Adverse Events with Efficacy in Patients With Small Cell Lung Cancer: a Second Analysis of Phase 3 IMpower133 Randomized Clinical Trials

**DOI:** 10.1101/2025.08.07.25333209

**Authors:** Sinbad Xia

## Abstract

**Introduction:** While immune-related adverse events (irAEs) are associated with improved outcomes for patients with non-small cell lung cancer (NSCLC), this relationship is unelucidated in small cell lung cancer (SCLC), where predictive biomarkers are critically needed.To evaluate the association between irAEs and overall survival (OS) in patients with extensive-stage SCLC (ES-SCLC) receiving first-line chemoimmunotherapy.

**Methods:** This secondary analysis utilized individual participant data from the IMpower133 study, a global, phase 3 randomized clinical trial that enrolled patients with previously untreated ES-SCLC receiving atezolizumab plus chemotherapy.

**Results:** The development of irAEs was not associated with improved OS. Patients who experienced an irAE had a median OS of 10.0 months compared to 9.77 months for patients without irAEs, a non-significant difference.

**Conclusions:** This finding challenges the established paradigm in NSCLC, underscores the distinct immunobiology of SCLC, and cautions against the extrapolation of biomarkers across thoracic malignancies, highlighting the urgent need for disease-specific biomarker discovery.

## Background

The emergence of immune-related adverse events (irAEs) has been established as a surrogate marker for clinical benefit in patients with metastatic non-small cell lung cancer (NSCLC) receiving PD-L1 inhibitors[1]. However, in the context of small cell lung cancer (SCLC), a malignancy characterized by distinct biological and clinical features[2], the relationship between irAEs and therapeutic outcomes has remained largely unelucidated[3]. While immune checkpoint inhibitors have been integrated into the standard-of-care for SCLC, a significant proportion of patients derive no clinical benefit, and robust predictive biomarkers are critically lacking[4]. To address this gap with a high level of evidence, this study performs a secondary analysis of data from a pivotal randomized controlled trial (RCT). By leveraging this robust dataset, we aim to systematically investigate the incidence and characteristics of irAEs in SCLC patients and to definitively assess their prognostic value, with the ultimate goal of informing clinical decision-making and patient stratification.

## Methods

Individual participant data from randomized phase Impower133 (ClinicalTrials.gov identifier: NCT02763579)[5] was utilized for this post-hoc analysis[6,7]. To evaluate the association between irAEs and efficacy in patients with small cell lung cancer (SCLC) using Impower 133 study.

### Statistical Analysis

Overall survival (OS) was defined as the time from randomization to death from any cause and was estimated using the Kaplan-Meier method. Survival distributions between patients who developed immune-related adverse events (irAEs) and those who did not were formally compared using a log-rank test. Median OS and the corresponding 95% confidence intervals (CIs) were calculated for each cohort. All statistical analyses were two-sided, and a p-value of less than 0.05 was considered to indicate statistical significance.

## Results

We assessed the association between irAE and overall survival (OS) in IMpower 133 study, irAE showed a slight increased risk of death (10 vs 9.77 months) versus patients without irAE (Fig 1).

**Figure 1.**
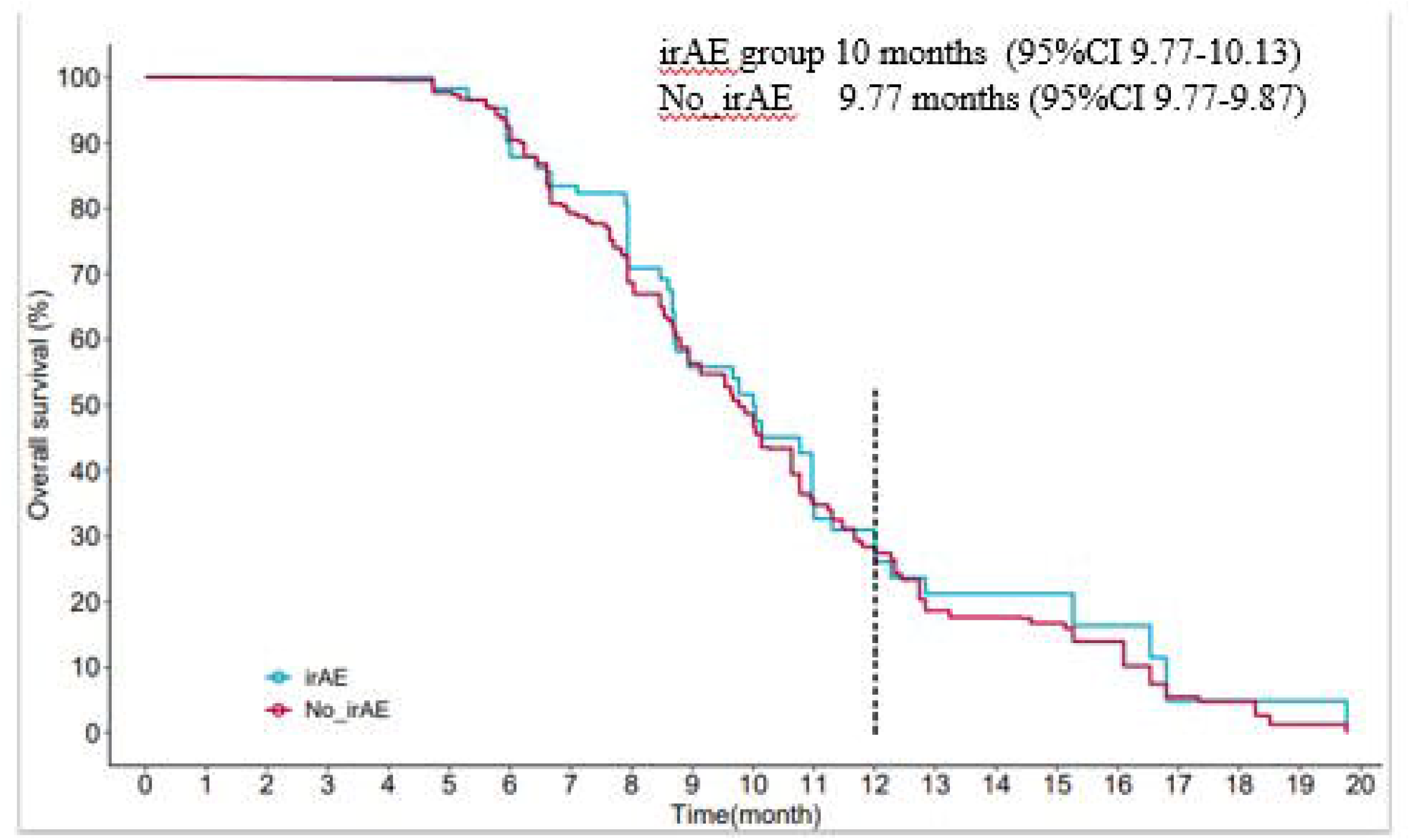
Kaplane Meier estimates of survival outcomes by irAE_in IMpower 133 study

## Discussion

This secondary analysis of the pivotal IMpower133 randomized controlled trial was conducted to elucidate the association between immune-related adverse events (irAEs) and clinical outcomes in patients with extensive-stage small cell lung cancer (ES-SCLC).

Contrary to the widely observed phenomenon in non-small cell lung cancer (NSCLC) and other solid tumors, our analysis did not confirm that the occurrence of irAEs serves as a surrogate marker for survival benefit in SCLC. In fact, our data revealed a non-significant trend towards a slightly shorter median overall survival (OS) in patients who developed irAEs compared to those who did not (10.0 vs 9.77 months). This unexpected finding challenges the prevailing hypothesis in the field.

The results we observed stand in stark contrast to findings in NSCLC, where irAEs are often interpreted as a reflection of effective anti-tumor immune activation. This disparity may be rooted in the unique tumor biology of SCLC[8], which is characterized by extreme aggressiveness, rapid doubling times, and a complex immunosuppressive tumor microenvironment. It is plausible that the systemic immune activation induced by immune checkpoint inhibitors, while sufficient to trigger irAEs, is inadequate to mount an effective and sustained cytotoxic effect against SCLC cells[9]. Alternatively, any modest anti-tumor activity may be overshadowed by the rapid pace of disease progression.

Furthermore, the confounding influence of treatment interruption or modification must be considered. Within the IMpower133 regimen (atezolizumab plus carboplatin/etoposide), patients experiencing moderate-to-severe irAEs may have required corticosteroid administration or even permanent discontinuation of immunotherapy or chemotherapy. For a disease as sensitive to treatment intensity and density as SCLC, any dose delay or reduction could have a direct negative impact on survival, potentially outweighing any theoretical benefit signaled by the irAE. As a post-hoc analysis, our inability to precisely capture and adjust for these detailed treatment modifications constitutes an inherent limitation.

The principal strength of this study lies in its use of data from a large, high-quality, Phase III randomized controlled trial, which provides a higher level of evidence than previous small, retrospective reports. However, the post-hoc nature of the analysis dictates its exploratory intent, and our lumped analysis of “any irAE” may have masked the heterogeneous effects of irAEs differing in type, severity, or timing of onset. This study’s limitation is its post-hoc design, creating a significant risk of bias. Crucial confounders, including treatment modifications or steroid use following an adverse event and inherent time-dependent biases, could distort the true association between irAEs and survival. Furthermore, the analysis pooled all irAEs regardless of type or severity and was not prospectively powered for this endpoint, meaning these findings must be considered exploratory and require validation in future dedicated studies.

In conclusion, this analysis of the IMpower133 data demonstrates no association between the occurrence of irAEs and improved survival in patients with ES-SCLC receiving first-line chemoimmunotherapy. This finding underscores the potential differences in immune response patterns between SCLC and NSCLC and cautions against the direct extrapolation of biomarkers from other tumor histologies to SCLC. Future studies should prospectively collect detailed data on irAEs and integrate translational research to deeply explore the unique mechanisms of immune activation and resistance in SCLC, with the aim of identifying truly applicable predictive biomarkers for this disease.

## Data Availability

All data produced are available online at https://vivli.org/

https://vivli.org/

## Funding

International Medical Association.

## Disclosure

All authors have declared no conflicts of interest.

## Acknowledgement

This presentation based on research using data from data contributors Roche that has been made available through Vivli, Inc. Vivli has not contributed to or approved, and is not in any way responsible for, the contents of this publication.

## Notes

### Competing Interest Statement

The authors have declared no competing interest.

### Funding Statement

This manuscript was produced support of International Medical Association.

### Author Declarations

This presentation based on research using data from data contributors Roche that has been made available through Vivli, Inc, https://vivli.org. Vivli has not contributed to or approved, and is not in any way responsible for, the contents of this publication.

### Summary of Updates

Revision summary: Add funding institution 8/23/2025

